# Accessibility and allocation of public parks and gardens during COVID-19 social distancing in England and Wales

**DOI:** 10.1101/2020.05.11.20098269

**Authors:** Niloofar Shoari, Majid Ezzati, Jill Baumgartner, Diego Malacarne, Daniela Fecht

## Abstract

Visiting parks and gardens may attenuate the adverse physical and mental health impacts of social distancing implemented to reduce the spread of COVID-19. We quantified access to public parks and gardens in urban areas of England and Wales, and the potential for park crowdedness during periods of high use. We combined data from the Office for National Statistics and Ordnance Survey to quantify (i) the number of parks within 500 and 1,000 metres of urban postcodes (i.e., availability), (ii) the distance of postcodes to the nearest park (i.e., accessibility), and (iii) per-capita space in each park for people living within 1,000m. We examined how these measures vary by city and share of homes that are flats. Around 25.4 million people can access public parks or gardens within a ten-minute walk, while 3.8 million residents live farther away; of these 21% are children and 13% are elderly. Areas with a higher share of flats on average are closer to a park but people living in these areas are potentially less able to meet social distancing requirements while in parks during periods of high use. Cities in England and Wales can provide residents with access to green space that enables outdoor exercise and play during social distancing. Cities aiming to facilitate social distancing while keeping public green spaces open might require implementing measures such as dedicated park times for different age groups or entry allocation systems that, combined with smartphone apps or drones, can monitor and manage the total number of people using the park.

## Introduction

On the 23^rd^ of March 2020, the UK government implemented a nationwide lockdown to reduce the transmission of COVID-19, which is in place until at least May. The measures require people to stay home at all times, except for shopping essentials or exercise, during which a minimum distance of two metres from others should be maintained. England’s Chief Medical Officer, in a daily media briefing said that “discussions about moving on to the next stage of the response to the pandemic would be premature until the nation has passed the peak number of deaths” [1], In the absence of a vaccine or widespread testing and contact tracing, UK residents are likely to face a prolonged period of social distancing, at best intermittently [2, 3],

While extended social distancing measures slow down COVID-19 transmission, they have potentially far-reaching impacts on public health and wellbeing [4, 5], Extended periods of confinement at home reduce physical activity, particularly among people with lower socio-economic status [6], and increase the risk of depression, anxiety, insomnia, and self-harm [7, 8], Lack of opportunities for outdoor exercise is particularly problematic for children as many parents work from home, and especially for disadvantaged communities living in overcrowded homes and inner-city flats without access to outdoor space or private gardens [9-12].

Public parks and gardens provide an opportunity for people to experience nature, engage in physical activity, and feel a sense of social belonging, and hence can at least partially mitigate the mental and physical harms of the lockdown [13-17]. Competing priorities - social distancing to reduce disease transmission while ensuring outdoor play and exercise in green space - has led to diverse policy and advocacy responses in the UK and other parts of the world, ranging from park closures to limiting park opening times and reducing services such as park benches, children’s play areas, and sports facilities [18, 19].

Data are needed on the availability and accessibility of public parks and gardens, so that together with social and public health considerations, we can effectively and safely use cities’ green space resources to mitigate the adverse impacts of lockdown and other social distancing measures. Here, we describe the availability and accessibility of publicly owned public parks and gardens for urban areas in England and Wales, and consider policy options for their allocation and use.

## Methods

### Data sources

Our analysis focused on urban areas in England and the three most populous cities in Wales (i.e., Cardiff, Swansea, and Newport). We focused on urban areas because those living in rural areas typically have private gardens and/or access to the countryside. The urban areas in England were defined using the Built-up Areas boundaries from the Office for National Statistics (ONS) [20] and in Wales using the boundary data provided by local authorities.

We used mid-2018 population estimates, by age group, from the ONS, available at Lower Super Output Area (LSOA), matched to postcodes using postcode headcount information from the 2011 UK Census as weights. Each urban postcode on average has 15 households. We aggregated populations into five age categories: children and young adolescents (0-16 years), young adults (16-30 years), middle-aged adults (31-50 years and 51-70 years), and the elderly (70+ years). We obtained information on type of accommodation (residential flat versus house) from the 2011 UK Census. We identified public parks and gardens using the OS MasterMap Open Greenspace Layer (version October 2019), which provides information on the location, physical boundary, and function of publicly accessible green space.

### Statistical Analysis

We used a geographic information system (GIS) to conduct the following analyses: First, we quantified the availability of public green space, defined as total number of parks and gardens within 500 and 1,000 metres circular buffers around all residential postcodes (referred to as availability). The 500 and 1,000 metres buffer sizes approximately represent five and ten minutes of walking for an adult, respectively [21], Second, we quantified the accessibility of green space, measured by spatial proximity, using the (Euclidean) distance between the centroid of each postcode and the nearest public park or garden. Third, to evaluate the feasibility of urban parks and gardens to facilitate social distancing, we quantified the per-capita space available in each public park and garden by dividing its total area by the population size, weighted based on the number of parks within a 1,000 metres circular buffer around each postcode. For example, if a given postcode had three parks available within its 1,000 metres buffer, 33% of population was assigned to each park. We used this measure as an indicator of the possible crowdedness of the park or garden. A minimum space of four square meters per person is required to maintain a distance of at least two metres if people were spread evenly within the park. If people concentrate in certain areas of the park, such as paths, the requirement is higher. We report the number of people, stratified by age group, in different ranges of per-capita space. Finally, we examined accessibility and mean per-capita space of parks and gardens in relation to the proportion of homes in an LSOA that are flats, where a higher proportion of flats indicates greater reliance on public parks and gardens for green space access [12].

We conducted the analyses in ArcMap v.10.5.1 (ESRI Ltd, Redlands, California) and R Statistical Software (Version 1.2.5001).

## Results

Our analysis covered 537,713 urban postcodes in England and Wales, with a total population of over 29 million. Of these, ~6.2 million (21%) were children and young adolescents (0-16 years), ~5.9 million (20%) young adults (16-30 years), ~14.3 million (49%) middle-aged adults (31-50 years and 51-70 years), and 2.9 million (10%) 70+ years of age. There were a total of 4,155 public parks and gardens in urban areas in England and Wales.

### Availability and accessibility of public parks and gardens

There is on average one (standard deviation [SD] 1.2) public park and garden available within 500 metres of the urban postcodes in England and Wales, and three (SD 2.8) public parks and garden within 1,000 metres. Forty-three percent of postcodes in England and Wales do not have any public green space within 500 metres, 34% have one, and 23% have two or more. Fourteen percent and 22% of postcodes have either no park or one park, respectively, within 1,000 metres whereas 63% of postcodes have at least two parks and gardens.

Urban residents in England and Wales, on average, live 557 metres away from their closest park or garden. Ten percent of the population (2.8 million) has at least one park in the immediate vicinity of their residence (< 100 metres), 59% (14.4 million) within a 500 metres, 28% (8.3 million) between 500 to 1,000 metres, and 13% (3.8 million) live more than 1,000 metres from a public park or garden.

There are substantial differences in distance to public green space between cities (Fig 1). Bristol, Liverpool, and London have the best accessibility with median distances of 281 metres, 284 metres, and 322 metres, respectively. By comparison, Newport (median distance of 673 metres), Swansea (611 metres), Coventry (522 metres), and Leeds (518 metres) are cities with the least accessibility. In terms of the percentage of population, −5% of people in Swansea and Coventry and 7% in Leeds and Manchester have a public park or garden in the immediate vicinity (< 100 metres) of their residence compared with 15% in Bristol, 14% in Liverpool and 12% in London (Fig 2). The former four cities also have the highest percentages of population with a park located more than 1,000 metres from their residence, ranging from 15% in Manchester to 30% in Swansea, compared with just 0.2% in Bristol, 1% in Liverpool, and 4% in London.

**Fig 1.**
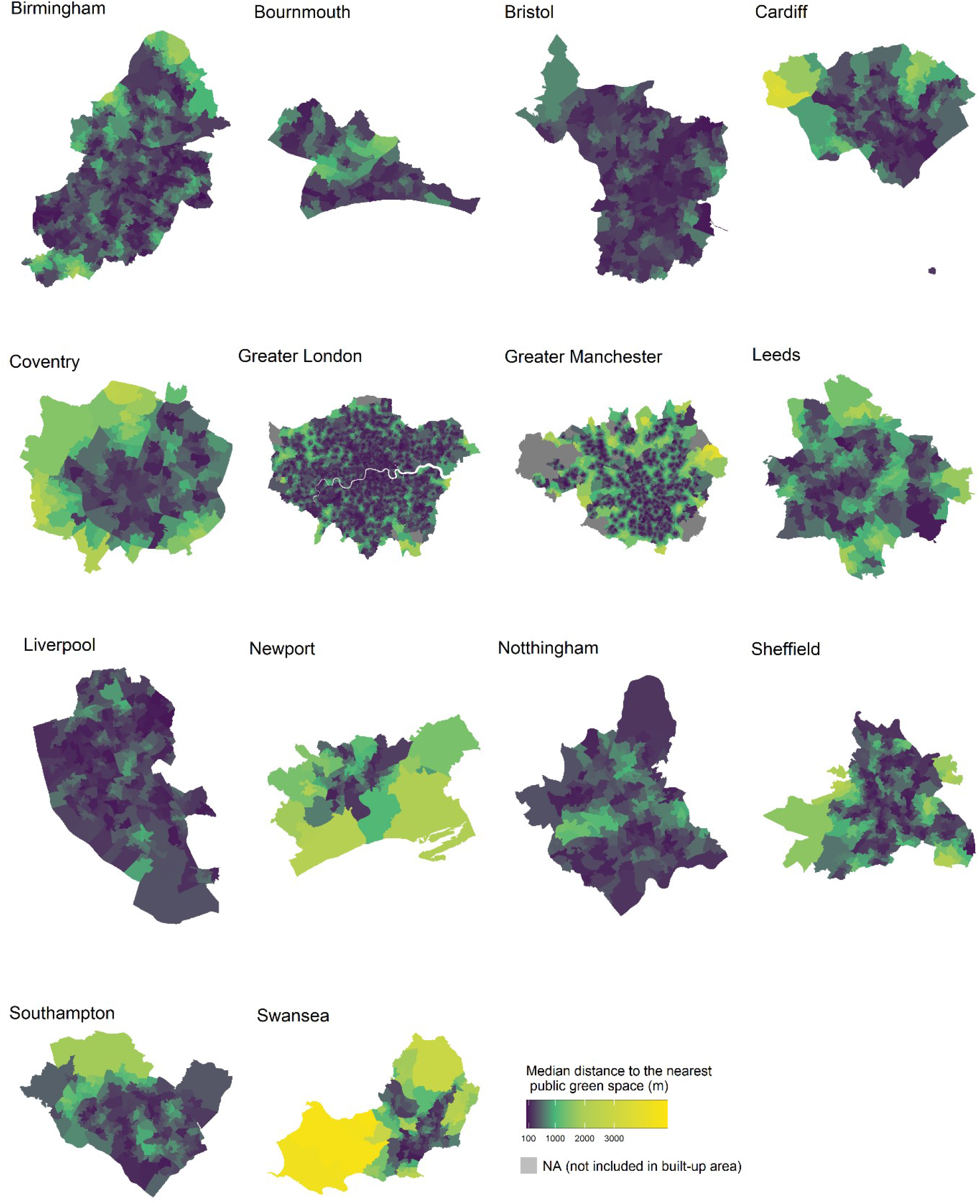
Median distance between home postcode and the nearest public park or garden in Lower Super Output Area (LSOA) for 14 cities in England and Wales. Postcodes were into LSOAs for presentation because there are a very large number of postcodes.

**Fig 2.**
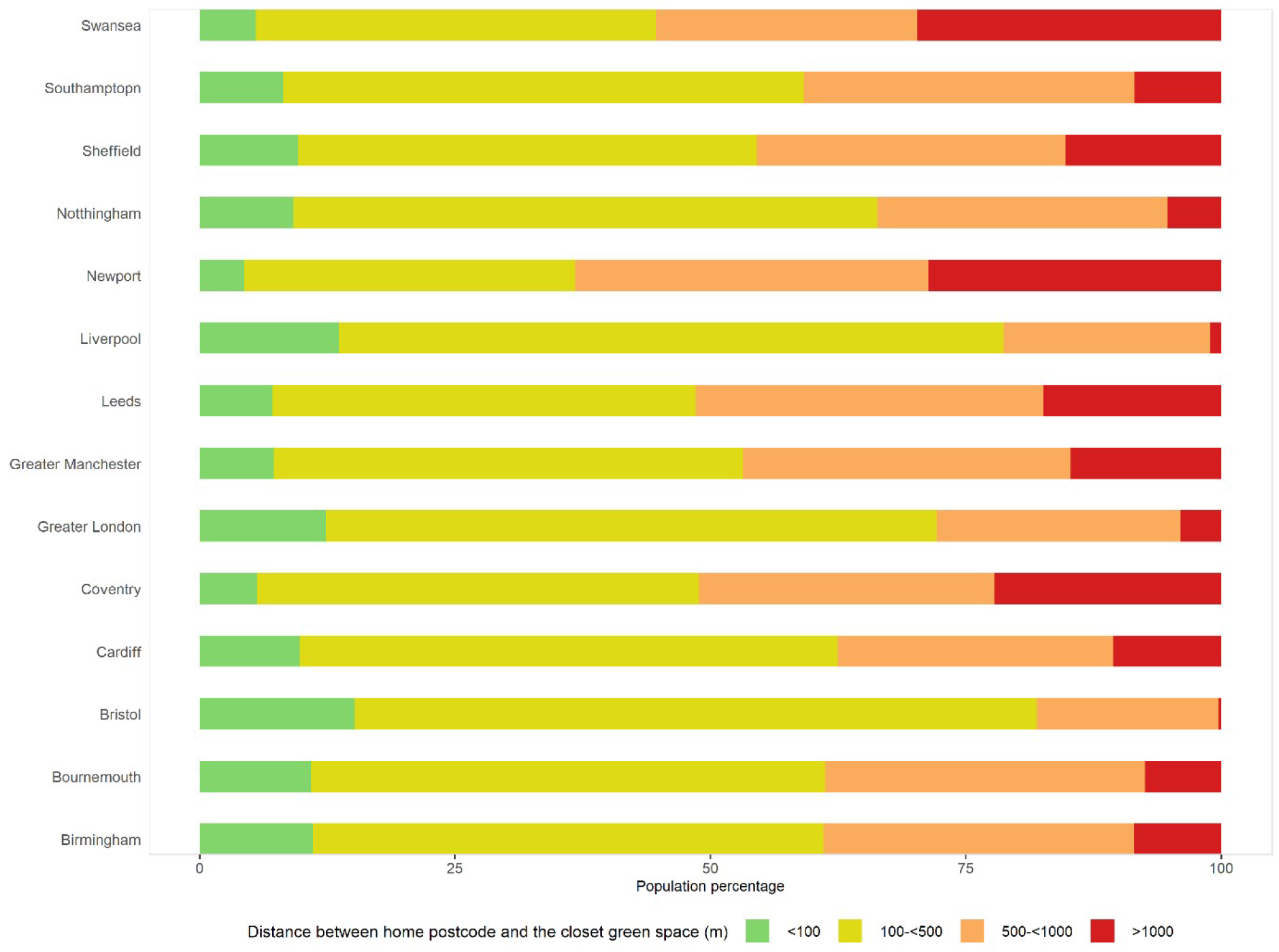
Percent of population in categories of distance to the closest park or garden for 14 cities in England and Wales.

### Can public parks and gardens facilitate social distancing?

Fig 3 shows available green space per-capita for all people (25.4 million) living within 1,000 metres of public parks and gardens in urban areas of England and Wales. At the extreme, if urban residents were all to visit their closest park at the same time, 50% of parks (2,071) would be unable to maintain the minimum social distancing space of four square metres per person, even if the entire park space was used. If each person visits the park for an hour over an eight-hour window (i.e. 12.5% of population visiting), an additional 39% of parks and gardens (89% total) would enable social distancing as long as people were equally spread out in the park.

**Fig 3.**
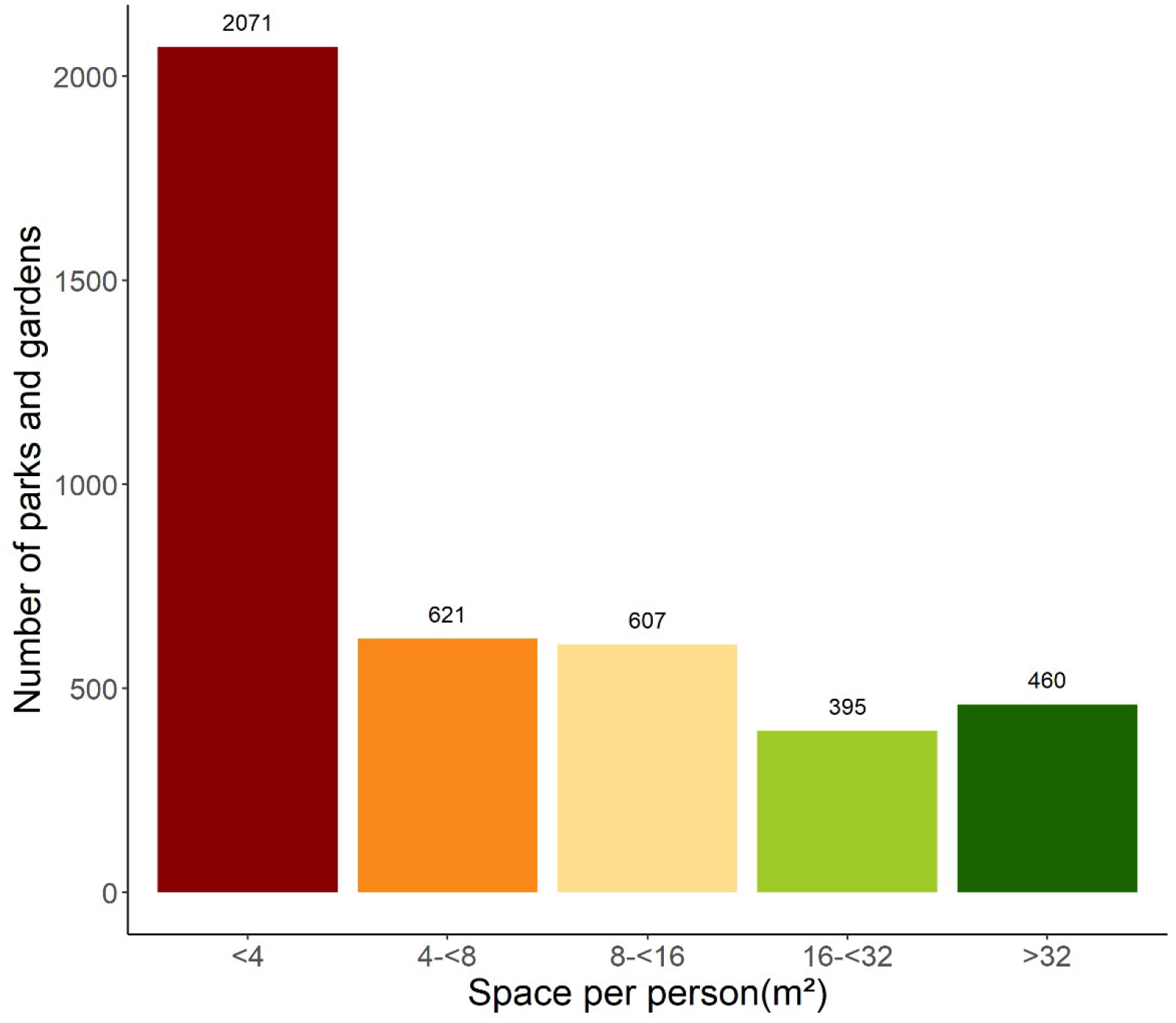
Distribution of space per person for public parks and garden in England and Wales.

For over 11 million of urbanites in England and Wales who have access to parks or gardens within 1,000 metres from their homes, there is a risk of going to a park (if all residents use the park simultaneously) that is so overcrowded that it disrupts social distancing (Fig 4). Of this group, 21% are children and young adolescents, and 9% are elderly. Around 3.8 million people do not have park or garden within 1,000 metres of their homes, of which 21% and 13% are younger than 16 years and 70 years and older, respectively.

**Fig 4.**
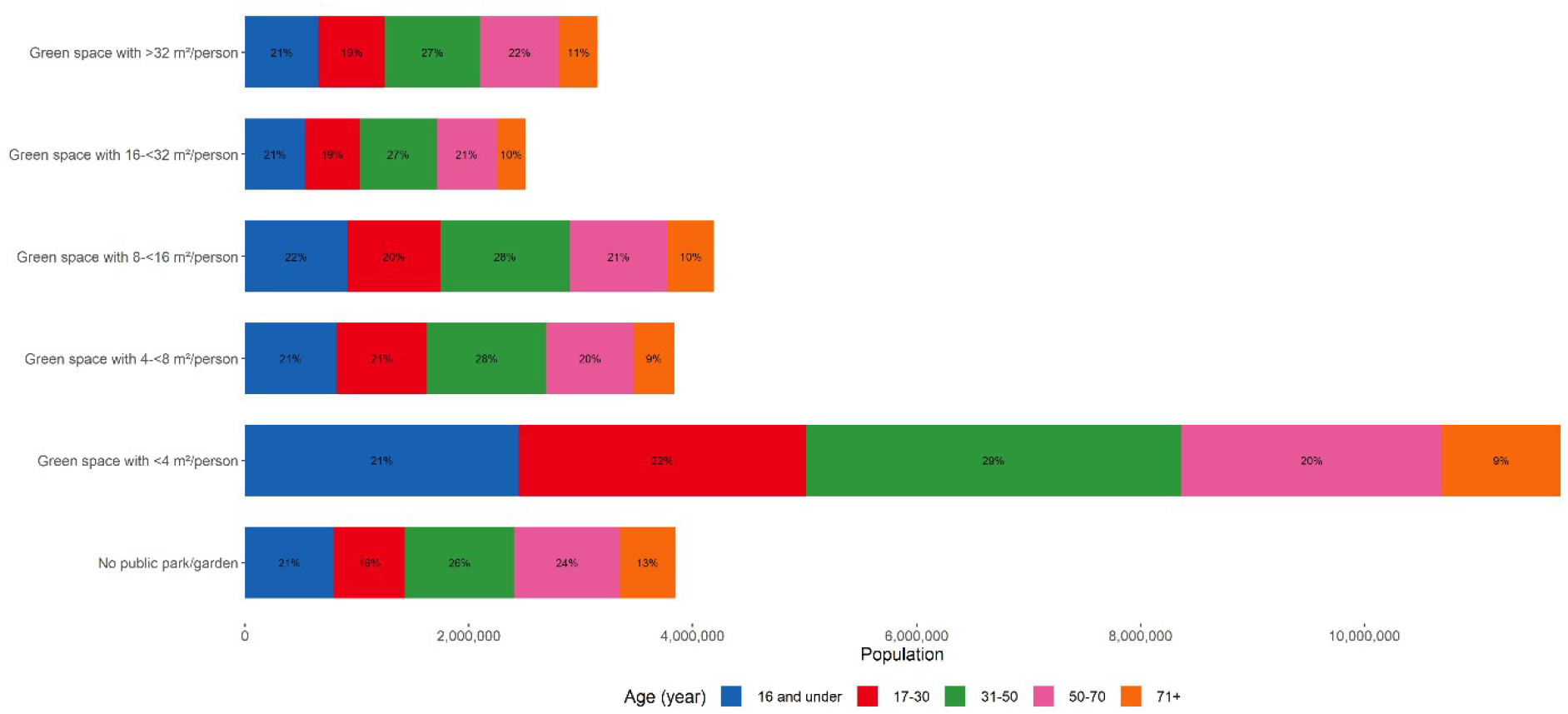
Number of people in each category of park space availability and age group. Numbers refer to the people living within 1,000 metres any park or garden.

LSOAs with a higher share of flats generally have a better accessibility to parks based on distance (Fig 5a), though parks in these LSOAs are more likely to be overcrowded if used by all residents (Fig 5b). For example, residents in the highest quantile of share of flats can reach a park or garden within 278 metres (the median) but the median space available in all parks in their LSOA is as small as 3.5 square metres per person.

**Fig 5.**
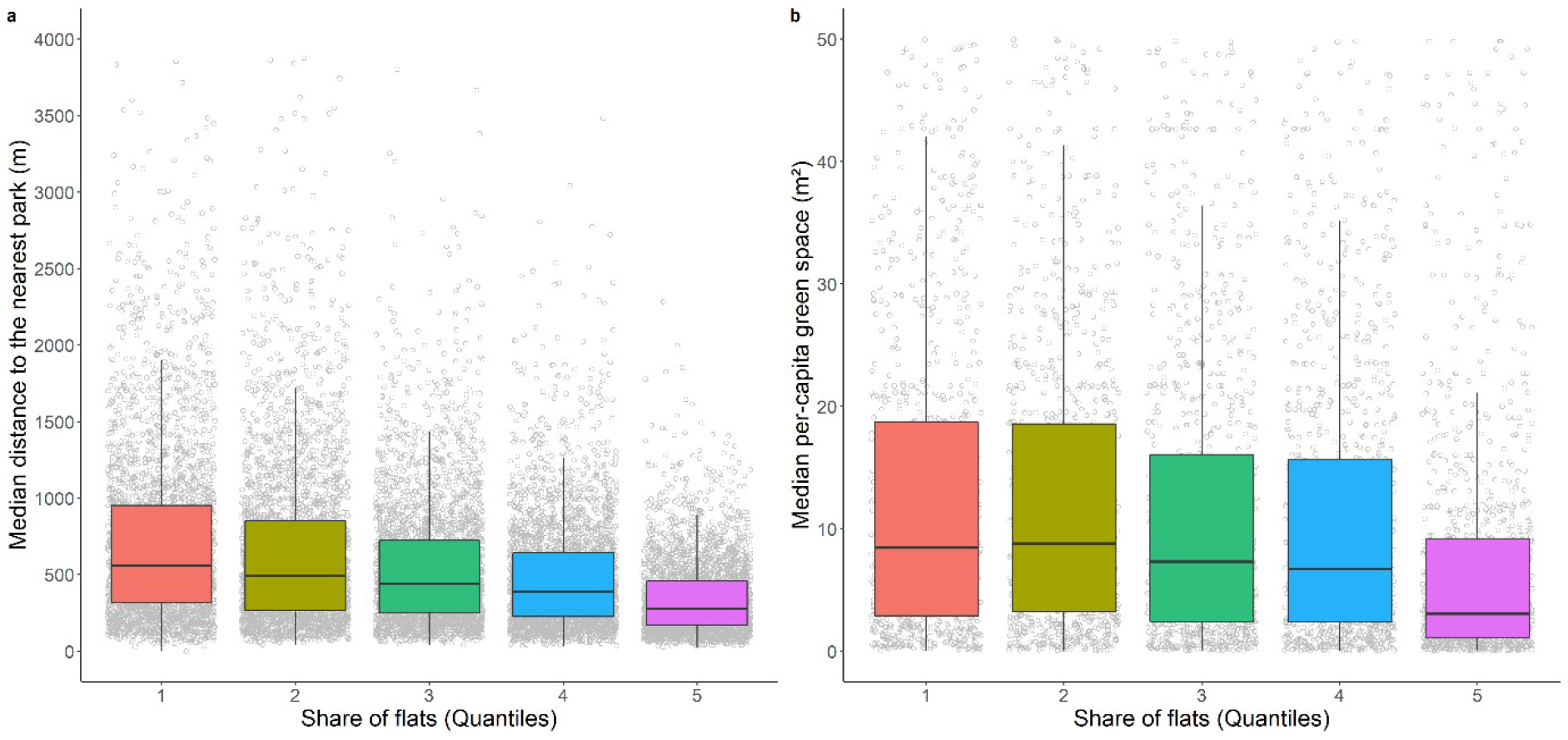
a) Median distance (m) to the nearest park or garden by quintiles of share of flats in Lower Super Output Area; and b) Median per-capita space (m^2^) by share of flats in Lower Super Output Area (quantiles), where 1 is the smallest proportion of flats and 5 is the highest proportion of flats. In Fig 5.b, the y-axis was delimited to 0 m^2^ and 50 m^2^ for the purpose of a better presentation.

## Discussion

Cities in England and Wales have green space assets that provide opportunities for outdoor exercise and play during lockdowns and other social distancing periods, but there are bottlenecks for some urbanites. Specifically, 13% of residents live more than 1,000 metres (-ten minutes walk) from their nearest public park or garden. Vising some parks and gardens, if used simultaneously by other residents, would not meet social distancing requirements.

The main strength of our study is to provide data on green space availability and accessibility that can inform options to keep public parks and gardens open and safe. A limitation of our work is that we did not consider other spaces (e.g. national parks and woodlands) although these are usually not located in urban areas. Further, our analysis could provide more detailed insights on crowding if there was information on time preferences for exercise and play, and attributes of parks beyond their amenities and services, which are closed during lockdowns, that attract specific people [22].

Public parks and gardens provide urban residents of all ages with access to outdoor green space and a place to exercise, which can reduce stress and improve mental and physical health [23, 24]. As social distancing measures continue into the spring and potentially longer, local and national governments will have to balance the access to outdoor green space with reducing the risk of transmission, particularly in densely populated areas. The Paris model of closing all parks reduces social contact in public spaces [25], but can have serious mental and physical health implications. Further, these more extreme measures may cause other outdoor spaces to become more crowded, risk engendering non-compliance with social distancing regulations, and can create tensions among residents and with officials that enforce the regulations.

An alternative policy is restricting access to high-risk areas (e.g., playgrounds and sports facilities) while keeping trails and open spaces accessible in a way that maintains social distancing. For example, parks can limit the number of people accessing based on park size and population density in the surrounding area [26], Dedicated park access times for different age groups or different activities could serve to both maintain social distancing and facilitate access for more vulnerable groups. Examples include specific times for families and the elderly and for walkers versus runners and cyclists. Alternatively, officials could manage utilisation, either based on weekly data to inform the community so that they can better spread their visits over the park’s opening times, or dynamically using smartphone or drone data to monitor crowdedness and communicate this information to residents.

Finally, given the extreme nature of the pandemic as a social and public health crisis, cities should complement public parks and gardens with other resources to lessen the adverse impacts of lockdown and social distancing. In Boston, Minneapolis and Oakland in United States, cities closed streets to vehicles to increase space for pedestrian and cyclists [27], Similarly, in the UK, some local authorities in London, Manchester, and Brighton are restricting driving on certain roads to separate walkers from runners and cyclists [28], Coordinating such an initiative would allow for longer routes and safer activities, provide alternative spaces for different activities (e.g., adult cyclists versus playing/running children), and potentially reduce congestion in parks. Opening up school green land, private parkland and golf courses to the public can provide additional space for exercising while maintaining social distancing [9], For example, Dulwich College and Dulwich Prep in London have opened up sections of their land to the public.

While a great deal of our attention in the early months of the pandemic is on supressing or stopping transmission, the strategies for achieving this can have detrimental impacts on health and wellbeing. Public parks and gardens are an important public health asset that can effectively help urban population to sustain their health and wellbeing, and should actively and effectively be used to do so.

## Data Availability

All input data is freely available from data providers as indicated in the manuscript. Derived data is available upon request from the authors.

## Declaration of interests

All authors have no conflicts of interests to declare.

## Acknowledgement

This work is supported by the Pathways to Equitable Healthy Cities grant from the Wellcome Trust [209376/Z/17/Z].

## Notes

### Competing Interest Statement

The authors have declared no competing interest.

### Funding Statement

This work is supported by the Pathways to Equitable Healthy Cities grant from the Wellcome Trust [209376/Z/17/Z]. The funder did not have any input in study design; in the collection, analysis and interpretation of data; in the writing of the report; and in the decision to submit the paper for publications.

